# Measures for infection prevention and control of SARS-CoV-2 in Belgian schools between December 2020 and June 2021: a prospective cohort study

**DOI:** 10.1101/2022.04.12.22273722

**Authors:** Milena Callies, Ines Kabouche, Isabelle Desombere, Joanna Merckx, Mathieu Roelants, Melissa Vermeulen, Els Duysburgh

**Author notes:** These authors contributed equally to this article and share first authorship (Milena Callies, Ines Kabouche).

## Abstract

**Introduction:** As the role of school-aged children was unclear at the onset of the COVID-19 pandemic, public health authorities recommended to implement infection prevention and control (IPC) measures in school settings. Few studies evaluated the implementation of these measures and their effect on SARS-CoV-2 infection rates among pupils and staff.

**Aim:** To describe the implementation of IPC measures in Belgian primary and secondary schools and assess its relation to the prevalence of anti-SARS-CoV-2 antibodies among pupils and staff.

**Methods:** We conducted a prospective cohort study in a representative sample of primary and secondary schools in Belgium. Implementation of IPC measures in schools was assessed using an online questionnaire. Saliva samples were collected from pupils and staff to determine the SARS-CoV-2 seroprevalence.

**Results:** A variety of IPC measures (ventilation, hygiene and physical distancing) was implemented by more than 60% of primary and secondary schools with most attention for hygiene measures. Almost no differences in implementation coverage were observed between primary and secondary schools or the Dutch and French language network. Poor implementation of IPC measures was associated with an increased anti-SARS-CoV-2 antibody prevalence among pupils from 8.6% (95% CI: 4.5 - 16.6) to 16.7% (95% CI: 10.2 - 27.4) and staff from 11.5% (95% CI: 8.1 - 16.4) to 17.6% (95% CI: 11.5 - 27.0). This association was statistically significant for all IPC measures and pupils and staff combined.

**Conclusion:** Belgian schools were relatively compliant with recommended IPC measures at the school level. Poor implementation of IPC measures was associated with higher SARS-CoV-2 seroprevalence among pupils and staff.

**Trial registration number:** Trial registration number: NCT04613817

## Introduction

In December 2019, a new coronavirus (SARS-CoV-2) was identified in Wuhan (China) causing high morbidity and mortality in the adult population which in turn led to a worldwide overload of healthcare resources. By January 2020, the World Health Organization (WHO) declared that the disease caused by the virus, COVID-19, could be characterized as a pandemic (1). Since then, international efforts have been made to reduce infection rates of SARS-CoV-2 and the burden on healthcare systems.

To control the spread of SARS-CoV-2, national and regional authorities implemented SARS-CoV-2 infection prevention and control (IPC) measures in various societal settings. Because the role of school-aged children in the transmission dynamics was unclear, IPC measures against COVID-19 were implemented in schools from an early stage (2). The assumption that school-aged children play a major role in maintaining the epidemic, as is the case for seasonal influenza, made many governments implement drastic measures such as school closures and switch to remote learning (3–5). The United Nations Educational, Scientific and Cultural Organization (UNESCO) reported that 190 countries closed their schools by April 2020 which affected 90% of the world’s school-aged population (6,7). The implementation of these measures was criticized as no evidence was available on the SARS-CoV-2 transmission among children and youth and the fact that these measures could cause additional mental and socio-economic problems among school-aged children (3). In response, many European countries reopened their schools and implemented a variety of IPC measures at school level with the objective to create an as safe as possible environment for in-person education. However, the implementation of these IPC measures differed nationally, regionally and across educational level (2).

International organizations (WHO, UNESCO, UNICEF, IFRC) and national public health authorities recommended a broad range of COVID-19 IPC measures for primary and secondary educational settings (8). The classification of these measures changed over time and differed across organizations and national entities (8–12). For example; the World Health Organisation (WHO) defined the following categories of measures: 1. personal protection, 2. environmental, 3. physical distancing, and 4. screening (8). Yet, a scoping review by *Krishnaratne et al*. defined other categories: 1. organizational measures, 2. structural/environmental measures, and 3. surveillance and response measures (2).

In Belgium, several IPC measures were recommended in schools which can be classified into the above-mentioned WHO categories. From mid-March until May 2020, remote learning was mandatory for all schools. Schools reopened for in-person teaching in May 2020 while implementing IPC measures such as hand and respiratory hygiene, ventilation and physical distancing measures and introducing an intensive contact tracing programme. Furthermore, the last four grades of secondary school (ages 14-17) were required to organize 50% of classes online in order to limit physical presence at school. Masking was compulsory for all primary and secondary school staff and all pupils from secondary schools (13).

Studies examining the impact of IPC measures in schools are limited (14). Design and analysis issues apply, making it difficult to evaluate the impact of individual or combined measures (15). One study estimated that the combination of reduced class density, transmission mitigation measures (use of masks, desk shields, frequent surface cleaning, outdoor instruction), and early identification of active infections would reduce SARS-CoV-2 prevalence (16). Additionally, a study in Barcelona, Spain, found that transmission rates of SARS-CoV-2 among children were lower in summer schools applying IPC measures such as contact bubbles, hand hygiene, masks and outdoor activities, compared to the general population (17). Similar findings were observed in primary and secondary schools in Switzerland (18). Despite the limited number of studies, most agree that the implementation of IPC measures in schools is associated with lower SARS-CoV-2 transmission rates compared to other settings such as households or the community (17,18). However, given the potential impact of IPC measures on pupil’s lives, a thorough evaluation of their impact on SARS-CoV-2 transmission in school environments is desired.

To our knowledge, no information has yet been published on the implementation of IPC measures against SARS-CoV-2 transmission in Belgian schools. The objective of this paper is to document the implementation of IPC measures in Belgian primary and secondary schools and to investigate its relation with the prevalence of anti-SARS-CoV-2 antibodies among pupils and staff.

## Methods

This analysis is based on data collected through a country-wide representative longitudinal seroprevalence study on SARS-CoV-2 antibodies among Belgian pupils and school staff during the school year 2020 – 2021 (19).

### Study design

Data on implementation of IPC measures were collected from schools at the start of the study in December 2020/January 2021, and again in March and May/June 2021. Saliva samples were taken from pupils and staff to determine the prevalence of anti-SARS-CoV-2 antibodies.

### Study population and recruitment

Schools, pupils and staff were recruited using a two-stage randomized cluster design with proportional allocation by province and sociodemographic background. In the first stage, 41 clusters were identified in which one primary school and one secondary school were selected at random from a list of all Belgian schools providing general education. Clusters were allocated per province proportional to the child population on January 1st 2020 (20). In the second stage a convenience sample of 20 pupils and 10 staff was recruited in each school. Inclusion criteria were being a pupil attending the 2^nd^ or 3^rd^ grade (ages 7 to 9 years) of primary school, or a pupil attending the 2^nd^ grade (ages 13 to 14 years) of secondary school, or a staff member in contact with eligible pupils.

### Sample size

A sample size of 800 pupils and 400 staff at each school level (primary and secondary schools) was calculated to estimate the seroprevalence with a margin of error of 2.3% for pupils and 3.0% for staff, assuming a seroprevalence of 6% among pupils and 10% among staff and a cluster design effect of two. All participants were recruited during the first data collection period (Dec 2020/Jan 2021).

### Data collection

The local study coordinator – a staff member of the participating school – collected the data on implemented IPC measures via a secured online questionnaire using the ‘LimeSurvey’ platform (LimeSurvey version 3.22.24+2000630; see Supplement I). The questionnaire was available in Dutch and French. The list of IPC measures assessed in the questionnaire was based on recommendations of the Belgian education authorities announced in November 2020 (21,22). These recommendations included guidelines on school closures, school and classroom ventilation, personal and environmental hygiene and physical distancing. The baseline questionnaire provided information regarding the period between the reopening of schools (May 2020) and the first data collection period (Dec 2020/Jan 2021). The follow-up questionnaires provided information regarding the period between the previous and present data collection period.

During each data collection period, the prevalence of anti-SARS-CoV-2 antibodies among pupils and staff of participating schools was assessed using saliva samples. These samples were self-collected via an Oracol device (Oracol, Malvern Medical Developments, UK) under the supervision of a trained nurse. Semi-quantitative levels of anti-RBD (Receptor Binding Domain) IgG were determined in each sample at the Immunology Laboratory of Sciensano (Public Health Belgium) using the WANTAI SARS-CoV-2 IgG ELISA (Quantitative) (Beijing, Wantai Bio-Pharm, China, cat n° WS-1396) customized for saliva samples using an in house protocol. Assay performance and a specificity-optimized cut-off of >1.5 signal-to-noise ratio for anti-RBD IgG positivity in saliva was evaluated using receiver operating characteristic analyses. Using this cut-off, the specificity of the test was 96.7% and 96.5% and the sensitivity 95.1% and 80.0% for adults and children, respectively.

### Implementation of IPC measures

IPC measures were grouped according to the target age as: (1) IPC measures applied in both, primary and secondary schools, (2) IPC measures applied in primary schools only, and (3) IPC measures applied in secondary schools only. Group 1 was further divided in four subcategories: (1) school/class closures, (2) ventilation measures, (3) hygiene measures (environmental & personal), and (4) physical distancing measures (Table 1). We based this classification on the one in the first ECDC technical report on schools with an additional subcategory for ventilation measures (14).

**Table 1:** Number and percentage of Belgian primary and secondary schools that implemented IPC measures^1^ during three data collection periods (Dec 2020/Jan 2021, March 2021 and May/June 2021)^2^.

| Specific IPC measure | Primary schools |  |  | Secondary schools |  |  |
| --- | --- | --- | --- | --- | --- | --- |
|  | M0<br>N=43<br>n (%) | M3<br>N = 44<br>n (%) | M6<br>N = 43<br>n (%) | M0<br>N=40<br>n (%) | M3<br>N=38<br>n (%) | M6<br>N=39<br>n (%) |
| **IPC measures applied in both primary and secondary schools (16 measures)** |  |  |  |  |  |  |
| Closures (2 measures) |  |  |  |  |  |  |
| School closure outside holiday breaks | 1 (2) | 3 (7) | 7 (16) | 0 (0) | 3 (8) | 1 (3) |
| Classes suspended | 23 (53) | 19 (43) | 18 (42) | 22 (55) | 13 (34) | 10 (26) |
| Ventilation measures (4 measures) |  |  |  |  |  |  |
| Classrooms have a CO<sub>2</sub> detector | 9 (21) | / | 11 (26) | 3 (8) | / | 4 (10) |
| School has and uses a ventilation system | 9 (21) | / | 4 (9) | 5 (13) | / | 5 (13) |
| Teachers are encouraged to ventilate class rooms regularly | 41 (95) | 42 (95) | 40 (93) | 39 (98) | 38 (100) | 35 (90) |
| Classes take place outside as much as possible | 3 (7) | / | 5 (12) | 2 (5) | / | 6 (16) |
| Hygiene measures (environmental & personal) (5 measures) |  |  |  |  |  |  |
| Classrooms are cleaned regularly and more frequently than previous school years | 26 (60) | 31 (70) | 31 (72) | 27 (68) | 32 (84) | 29 (74) |
| Staff rooms are cleaned regularly and more frequently than previous school years | 28 (65) | 33 (75) | 32 (74) | 25 (63) | 33 (87) | 29 (74) |
| Toilets are cleaned regularly and more frequently than previous school years | 31 (72) | 40 (91) | 37 (86) | 31 (78) | 38 (100) | 31 (79) |
| Surfaces that are touched regularly are disinfected daily | 33 (77) | 33 (75) | 33 (77) | 33 (83) | 30 (79) | 26 (67) |
| Alcohol gel (or additional possibilities to clean hands) is made available for pupils and staff | 39 (88) | 42 (95) | 40 (93) | 38 (95) | 37 (97) | 37 (95) |
| Physical distancing measures (5 measures) |  |  |  |  |  |  |
| Breaks are spread to decrease contact between different age groups | 12 (28) | 16 (36) | 13 (30) | 18 (45) | 17 (45) | 9 (23) |
| Number of staff is limited per room | 32 (74) | 35 (80) | 35 (81) | 33 (83) | 30 (79) | 29 (74) |
| Pupils have one fixed place in a fixed classroom | 25 (58) | 32 (73) | 31 (72) | 28 (70) | 30 (79) | 31 (79) |
| Teachers change between classrooms, not the pupils | 22 (51) | 31 (70) | 27 (63) | 28 (70) | 28 (74) | 28 (72) |
| Lunches are taken in the classroom. If this is not possible pupils have a fixed place in the dining area | 36 (84) | 42 (95) | 38 (88) | 34 (85) | 35 (92) | 30 (77) |
| **IPC measures applied in primary schools only (3 measures)** |  |  |  |  |  |  |
| Staff wear a mask if sufficient distance cannot be maintained | 34 (79) | 43 (98) | 39 (91) |  |  |  |
| Distance is kept during contacts between adults | 38 (88) | 39 (89) | 34 (79) |  |  |  |
| Distance is kept during contacts between staff and pupils | 24 (56) | 18 (41) | 24 (56) |  |  |  |
| **IPC measures applied in secondary schools only (2 measures)** |  |  |  |  |  |  |
| Secondary schools only: Staff and pupils always wear a mask inside |  |  |  | 37 (93) | 36 (95) | 33 (85) |
| Secondary schools only: Staff and pupils wear a mask outside unless they can keep sufficient distance |  |  |  | 36 (90) | 37 (97) | 33 (85) |

<sup>1</sup> All measures, except for closures, are scored on a scale ranging from 1 (not applied at all) to 5 (fully applied). Measures with a score of '4' or '5' are considered 'implemented' by the school.
<sup>2</sup> Schools who did answer to certain measures were considered to not apply these measures. The number of missing schools is shown in Table S2.
IPC: infection prevention and control; M0: data collection period at study month 0, 3 Dec 2020 - 28 Jan 2021; M3: data collection period at study month 3, 1 - 26 Mar 2021; M6: data collection period at study month 6, 17 May – 11 Jun 2021; n (%): absolute number of schools (percentage of schools) that implemented the measure; N: number of schools that completed the questionnaire.

For each data collection period, we reported the number and proportion of schools that implemented the IPC measure. For school and class closures, we reported the number and proportion of schools with at least one closure during the assessed period. The other IPC measures were assessed using a 5-point Likert scale ranging from 1 (the IPC measure was ‘not applied at all’) to 5 (the IPC measure was ‘fully applied’). For the analysis we considered measures with a score of 4 or 5 as ‘implemented’ and with a score from 1 to 3 as ‘not implemented’. When a school did not respond for a specific measure (Table S2, S3), we considered it as ‘not implemented’. The implementation of IPC measures was analysed at school level (primary versus secondary schools), data collection period (Dec 2020/Jan 2021, March 2021, May/Jun 2021) and language network level (French versus Dutch).

### Relation between IPC measures implementation and prevalence of anti-SARS-CoV-2 antibodies

We assessed the relation between the implementation of IPC measures and prevalence of anti-SARS-CoV-2 antibodies among pupils and staff in primary and secondary schools that provided an answer for at least 10 out of 14 measures included in this analysis. Measures related to school or class closures and measures not included in both, primary and secondary schools, were not included. Schools were classified according to their compliance with the implementation of IPC measure as poor, moderate or thorough. This classification was based on the sum of Likert scale scores (each ranging from 1 to 5) for individual IPC measures. Schools in the upper quartile were designated as ‘thorough’, and schools in the lower quartile as ‘poor’ implementers. Schools with a sum of scores in between were considered ‘moderate’ implementers. Missing scores were replaced with the mean score from the remaining school for a maximum of 4 measures. The prevalence and 95% confidence interval (95% CI) of anti-SARS-CoV-2 antibodies among pupils and staff for the first data collection period was calculated in each group with generalized estimation equations to account for possible clustering of cases in schools. Generalized estimation equations for binomial outcomes with a log link function were used to assess if the implementation of IPC measures (poor, moderate, thorough) was associated with the prevalence of anti-SARS-CoV-2 antibodies, taking into account the community exposure (total reported cases in the school district 14 days before testing), socioeconomic tertile and language network (Dutch, French) of the school, type of school (primary, secondary), subject category (staff, pupil; when applicable), and school identification as the clustering variable with an exchangeable correlation structure. Results are expressed as an adjusted Relative Risk (aRR) with 95% CI. This analysis was done for all IPC measures together and for the IPC subcategories (ventilation, hygiene and physical distancing). This analysis is based on data from the first data collection period which is the most complete and correct assessment (for example, during the second period less ventilation measures were assessed and seroprevalence data from the third data collection period were impacted by vaccination among staff).

Data were analysed using SAS Enterprise Guide version 7.1 (SAS Institute Inc., Cary, NC, USA) and R version 4.0 (2021, R. Foundation for statistical computing, Vienna, Austria)

### Ethics approval

The study was approved by the Medical Ethics Committee of the University Hospital Ghent (reference: B6702020000744 - BC-08564). Written informed consent was obtained from all staff and parents of participating pupils before enrolment. Additionally, written informed assent was obtained from all pupils.

## Results

### Participating schools

We contacted 98 primary and 108 secondary schools of which 44 primary and 40 secondary schools agreed to participate. Of these 84 schools, 45 belonged to the Dutch and 39 to the French language network. All but one school (n = 83) completed the online questionnaire in Dec 2020/Jan 2021 (first data collection period) and 82 schools in March and May/Jun 2021.

Requirements for the assessment of the association between IPC measures and anti-SARS-CoV-2 antibody prevalence (at least 10 of 14 measures with an answer) were met by 81 schools (43 primary and 38 secondary). Among these, 21 (26%) schools were classified as implementing the measures thoroughly, 37 (46%) as moderately and 23 (28%) poorly. The mean prevalence of anti-SARS-CoV-2 antibodies for each group was calculated based on a total of 1,285 pupils (710 primary, 575 secondary school pupils) and 818 staff (432 primary, 386 secondary school staff).

### Implementation of IPC measures

Table 1 shows the number and proportion of schools that implemented each of the individual IPC measures during the three data collection periods. Similar implementation in primary and secondary schools was observed for most IPC measures. Between the reopening of schools in May 2020 and the last data collection period in May/Jun 2021, 13 (16%) schools were closed due to COVID-19. Hygiene measures were implemented most frequently, followed by physical distancing and ventilation. The implementation of individual IPC measures did not change substantially over the three data collection periods, but between March and May/Jun 2021, school closures were more frequent in primary than in secondary schools (Table 1).

Teachers were systematically encouraged to ventilate classrooms (93% of primary and 90% of secondary schools by May/Jun 2021), but a noticeable lower number of schools invested in the use of CO2 detectors (26% of primary and 10% of secondary schools by May/Jun 2021) or active ventilation systems (9% of primary and 13% of secondary schools by May/Jun 2021). Classes were rarely organized outdoors. Physical distancing measures were implemented in the majority of schools, except for the separation of age groups during breaks (30% of primary and 23% of secondary schools by May/Jun 2021). Findings from May/Jun 2021 show further that pupils from 72% of secondary schools stayed in the same classroom instead of changing classrooms as usual. Most hygiene measures were implemented by more than 75% of schools during the three data collection periods. Mask wearing among staff of primary and secondary schools and pupils of secondary schools was reported to be well implemented (more than 80% compliance) (Table 1).

Implementation of IPC measures was overall similar in both language networks (Table S1). An exception is the use of fixed classrooms for pupils, which was more common in the Dutch language network (89% of schools) compared to the French language network (59% of schools) in May/Jun 2021. For this measure, the opposite was noticed for teachers (49% Dutch and 82% French language network).

### Relation between IPC measures implementation and prevalence of anti-SARS-CoV-2 antibodies among pupils and staff

Figure 1 shows that, taking all IPC measures together, the prevalence of anti-SARS-CoV-2 antibodies increased from 8.6% (95% CI: 4.5 – 16.6) to 16.7% (95% CI: 10.2 – 27.4) among pupils and from 11.5% (95% CI: 8.1 -16.4) to 17.6% (95% CI: 11.5 – 27.0) among staff with poorer implementation of these measures. This association was statistically significant for pupils and staff combined (aRR 0.79, 95% CI 0.64 – 0.98, p = 0.03), meaning a decrease of being seropositive by 21% when implementation of IPC measures is thorough. However, the association was not statistically significant for pupils (aRR 0.77, 95% CI 0.53 - 1.10, p = 0.15) or staff (aRR 0.81, 95% CI 0.62 - 1.06, p = 0.12) separately.

**Figure 1:**
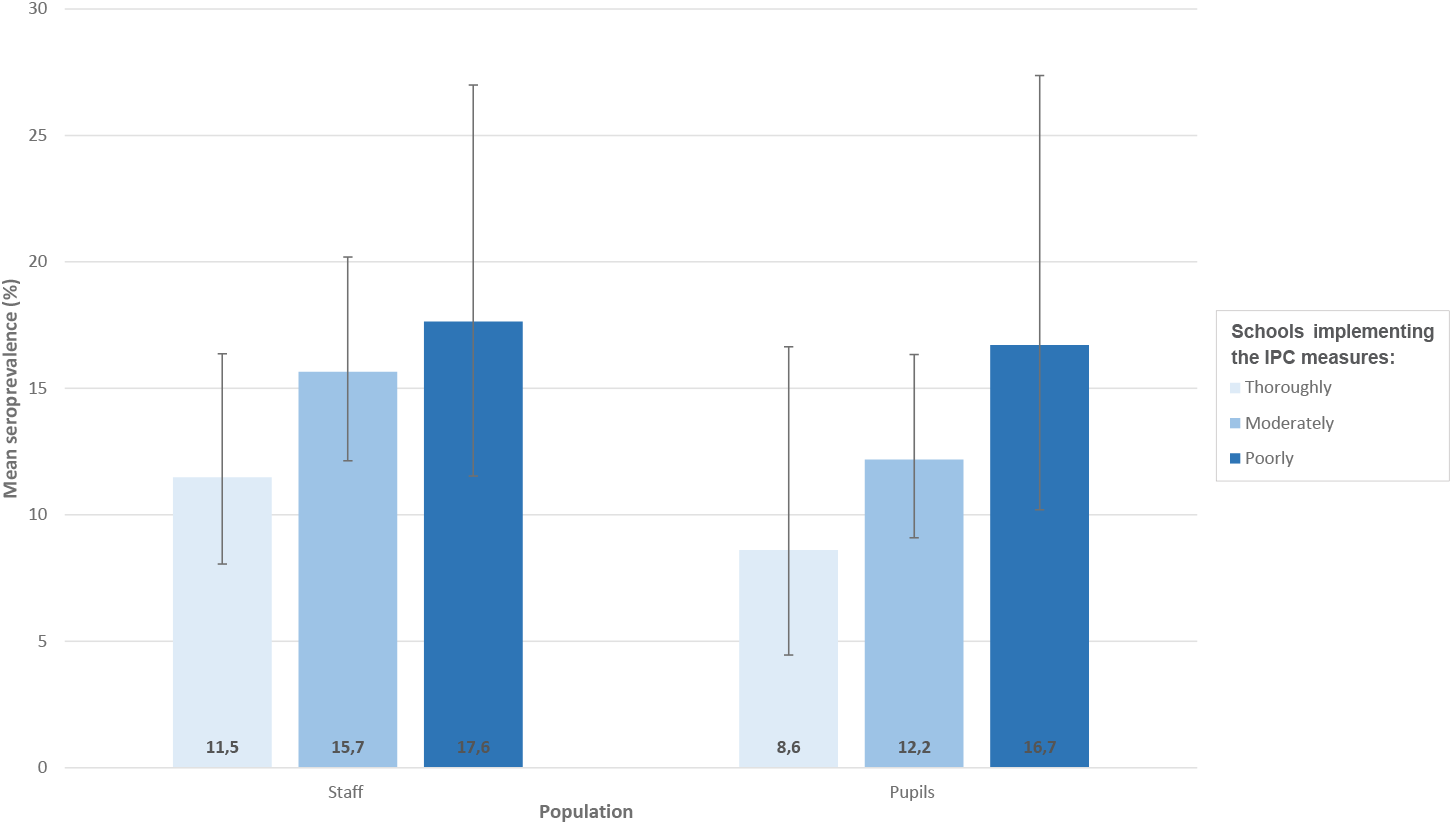
Prevalence of anti-SARS-CoV-2 antibodies among pupils and staff according to the degree of implementation of all school IPC measures, Belgian schools, Dec 2020/Jan 2021. The black lines indicate the upper and lower 95% confidence intervals; IPC: infection prevention and control.

When this analysis was repeated for each subcategory of measures (Figure 2), similar tends were observed meaning poorer implementation of IPC measures resulted in increased seroprevalence. However, none of them was found statistically significant (ventilation aRR 0.96, 95% CI 0.76 -1.22, p = 0.76; hygiene aRR 0.86, 95% CI 0.69 -1.07, p = 0.18; and physical distancing aRR 0.90, 95% CI 0.73 -1.12, p = 0.35).

**Figure 2:**
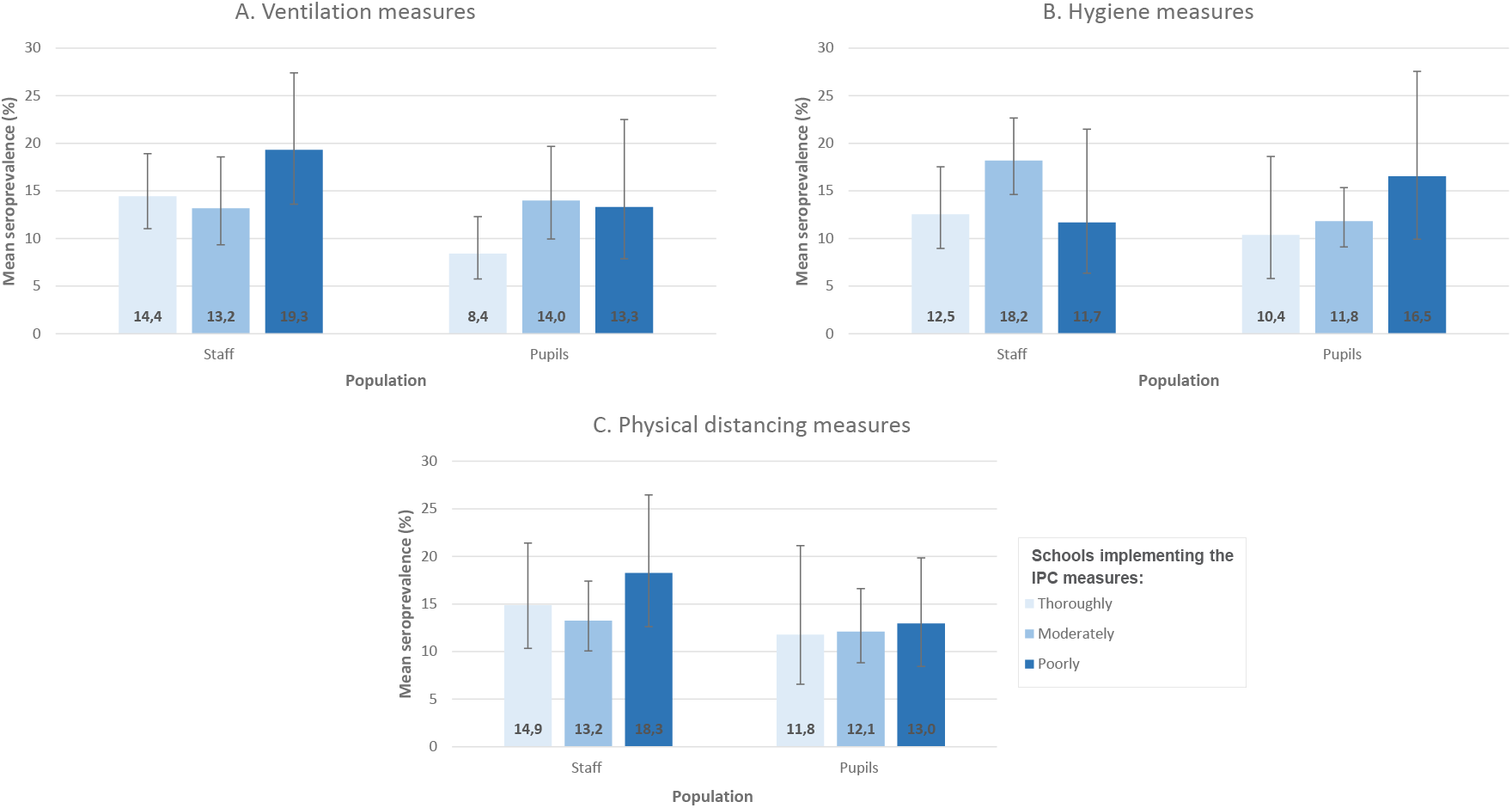
Prevalence of anti-SARS-CoV-2 antibodies among pupils and staff according to the degree of implementation of school ventilation (A), hygiene (B) and physical distancing (C) infection prevention and control measures, Belgian schools, Dec 2020/Jan 2021. The black lines indicate the upper and lower 95% confidence intervals; IPC: infection prevention and control

## Discussion

Apart from some ventilation and physical distancing measures, most IPC measures were implemented by more than 60% of primary and secondary schools. Implementation of IPC measures was quite similar in primary and secondary schools, but since the reopening in May 2020, more primary than secondary schools were closed. Schools that implemented less IPC measures usually had a higher seroprevalence of SARS-CoV-2 antibodies among pupils and staff, but this was only statistically significant for all measures combined in all subjects (pupils and staff). Despite observing comparable effect sizes in pupils and staff separately, these did not reach statistical significance, and neither were the trends for the three different IPC measures subcategories separately.

The number of school closures was higher in primary than in secondary schools. This might be due to the general recommendation and available infrastructure to organize distance learning in secondary, but not in primary schools. Secondary schools could thus easily adjust the amount of distance learning, while this option was not available for primary schools. The general recommendation for secondary schools in Belgium was to organize distance learning for half of the school population for grades 3 and higher (13). Although this recommendation did not apply to the secondary school pupils targeted in our study (2^nd^ grade), they limited the overall physical presence of pupils and staff in schools. Yet, the effect of school closures on the seroprevalence is less clear, largely because of the overlap with other IPC measures. A study from Sweden found that closures of higher grades in secondary schools did not necessarily have a large impact on SARS-CoV-2 infections (23).

Hygiene measures were well applied in most schools, possibly as a result of the widespread attention for hand hygiene, disinfection and cleaning from the onset of the pandemic (24), and because these measures are usually easy to implement. Ventilation measures were less applied, maybe because some of these require a higher financial investment as the installation of CO_2_ detectors or ventilation systems, while measures like passive ventilation may be challenging during winter. In the group of physical distancing measures, staggered break times by age group were less often implemented, which could be due to logistic and infrastructural challenges. In general, physical distancing measures were more frequently applied in schools from the Dutch language network which reflects differences in regional policies, school organization and regulations (13).

Mask wearing was reported to be almost fully implemented by primary and secondary school staff and by secondary school pupils. Although we could not assess the effect of mask wearing on SARS-CoV-2 prevalence in our study, findings in kindergarten and primary schools in Georgia (USA) suggest that this controls the transmission of SARS-CoV-2 in school settings (25). Also, a report on a SARS-CoV-2 outbreak in a primary school in California (USA) showed a higher SARS-CoV-2 transmission among pupils sitting closer to the teacher who did not wear a mask and was identified as the index case (26).

A statistically significant association was found in our study between the prevalence of anti-SARS-CoV-2 antibodies and the implementation of IPC measures in pupils and staff together but not separately. Since effect sizes, expressed as aRR, are of the same magnitude in both groups this is probably the result of a loss of statistical power. Compliance with recommendations regarding IPC measures might thus be beneficial for the total school population. Results for the different subcategories of IPC measures point in the same direction, but effect sizes are notably smaller and not statistically significant. Studies do suggest that ventilation measures might be more effective (25), but more likely a combination of measures is needed to reduce the transmission of SARS-CoV-2 as illustrated by the ‘Swiss-Cheese Model’ (27). A study in the USA for instance found that the risk associated with in-person teaching decreases when more IPC measures were implemented (28). Although a combination of measures subcategories would work best to prevent the spread of SARS-CoV-2, it is important to apply only those that would be most effective while taking into account the children’s educational needs and the staff’s general wellbeing.

A strength of our study is the random selection of a geographically and socially proportional sample of pupils and staff from Belgian schools. Also, this study is the first to assess the implementation of IPC measures in schools in relation to the prevalence of anti-SARS-CoV-2 antibodies among pupils and staff in Belgium. There are also limitations. First, the data on the implementation of IPC measures were self-reported which is prone to confirmation bias and misinterpretation. Secondly, data collection started in December 2020 and covered the preceding months which could result in recollection bias. Moreover, the first testing period covered a rather heterogeneous phase of the pandemic including periods of in-person teaching and the summer holiday in July and August 2020. Thirdly, individual characteristics of the participants were not included in our analyses. Finally, while representative for Belgian schools, the sample sizes at the school level are small and do not allow to quantify the attributable impact of individual IPC measures.

In conclusion, Belgian primary and secondary schools of both language networks complied relatively well with recommended IPC measures. A poor implementation of the IPC measures together and by ventilation, hygiene and physical distancing subcategory, showed an increase in the prevalence of anti-SARS-CoV-2 antibodies among pupils and staff in Belgian schools. This association was statistically significant for the IPC measures together for pupils and staff combined showing a decrease of being seropositive by 21% with thorough implementation. Despite observing comparable effect sizes in pupils and staff separately, these did not reach statistical significance, and neither were the trends for the three different IPC measures subcategories. This might be due to loss of statistical power.

## Data Availability

All data produced in the present study are available upon reasonable request to the authors.

## Conflict of interest

None

## Funding and acknowledgments

This study was funded by the Belgian government (COVID-19 SC_044A) through Sciensano, the Belgian institute of Public Health, Brussels, Belgium. Sciensano was involved in all stages of the study, from conception and implementation to analysis and reporting.

## Author’s contribution

Concept, design, protocol writing: J. Merckx, M. Roelants, E. Duysburgh.

Recruitment of participants and logistical coordination: M. Callies, I. Kabouche, M. Vermeulen.

Administrative, technical, or material support: I. Kabouche, M. Callies, E. Duysburgh, J. Merckx, M. Roelants, M. Vermeulen, I. Desombere.

Biological sample collection and transport: M. Callies, I. Kabouche.

Biological sample analysis: I. Desombere.

Epidemiological data collection: I. Kabouche, E. Duysburgh.

Epidemiological data cleaning and analysis: M. Roelants, M. Callies.

Drafting of manuscript: I. Kabouche, M. Callies.

Manuscript revision: E. Duysburgh, J. Merckx, M. Roelants, M. Vermeulen, I. Desombere.

Funding: E. Duysburgh.

Supervision: E. Duysburgh.

## Acknowledgements

All participants: Primary and secondary schools, school staff, pupils and their parents

Biological sample collection: All participating nurses

Biological sample transportation: Sciensano’s Sample Management (BBC transport services)

Biological sample analysis: Caroline Rodeghiero and Fabienne Jurion, expert technicians at the Department of Infectious Diseases in Humans, Immune Response, Sciensano, Brussels, Belgium

## Notes

### Competing Interest Statement

The authors have declared no competing interest.

### Clinical Protocols

https://www.sciensano.be/en/biblio/prevalence-and-incidence-antibodies-against-sars-cov-2-children-and-school-staff-measured-one-year

### Author Declarations

The Medical Ethics Committee of the University Hospital Ghent gave ethical approval for this work (reference: B6702020000744 - BC-08564).

## References

1. WHO Director-General’s opening remarks at the media briefing on COVID-19 - 11 March 2020 [Internet]. [cited 2021 Aug 5]. Available from: https://www.who.int/director-general/speeches/detail/who-director-general-s-opening-remarks-at-the-media-briefing-on-covid-19---11-march-2020

2. Krishnaratne S, Pfadenhauer LM, Coenen M, Geffert K, Jung-Sievers C, Klinger C, et al. Measures implemented in the school setting to contain the COVID‐19 pandemic: a rapid scoping review. Cochrane Database Syst Rev [Internet]. 2020 [cited 2021 Aug 5];(12). Available from: https://www.cochranelibrary.com/cdsr/doi/10.1002/14651858.CD013812/full

3. Esposito S, Cotugno N, Principi N. Comprehensive and safe school strategy during COVID-19 pandemic. Ital J Pediatr. 2021 Jan 9;47(1):6.

4. Esposito S, Principi N. School Closure During the Coronavirus Disease 2019 (COVID-19) Pandemic: An Effective Intervention at the Global Level? JAMA Pediatr. 2020 Oct 1;174(10):921–2.

5. Litvinova M, Liu Q-H, Kulikov ES, Ajelli M. Reactive school closure weakens the network of social interactions and reduces the spread of influenza. Proc Natl Acad Sci U S A. 2019 Jul 2;116(27):13174–81.

6. Yoon Y, Kim K-R, Park H, Kim S, Kim Y-J. Stepwise School Opening and an Impact on the Epidemiology of COVID-19 in the Children. J Korean Med Sci [Internet]. 2020 Nov 20 [cited 2021 Aug 6];35(46). Available from: https://synapse.koreamed.org/articles/1146252?viewtype=pubreader

7. https://plus.google.com/+UNESCO. Education: From disruption to recovery [Internet]. UNESCO. 2020 [cited 2021 Aug 6]. Available from: https://en.unesco.org/covid19/educationresponse

8. Considerations for school-related public health measures in the context of COVID-19 [Internet]. [cited 2021 Aug 5]. Available from: https://www.who.int/publications-detail-redirect/considerations-for-school-related-public-health-measures-in-the-context-of-covid-19

9. Key Messages and Actions for COVID-19 Prevention and Control in Schools. :13.

10. Melnick H, Darling-Hammond L. Reopening Schools in the Context of COVID-19: Health and Safety Guidelines From Other Countries. :13.

11. Bonell C, Melendez-Torres GJ, Viner RM, Rogers MB, Whitworth M, Rutter H, et al. An evidence-based theory of change for reducing SARS-CoV-2 transmission in reopened schools. Health Place. 2020 Jul 1;64:102398.

12. Johansen TB, Astrup E, Jore S, Nilssen H, Dahlberg BB, Klingenberg C, et al. Infection prevention guidelines and considerations for paediatric risk groups when reopening primary schools during COVID-19 pandemic, Norway, April 2020. Eurosurveillance. 2020 Jun 4;25(22):2000921.

13. Proesmans, Bloemen, Hancart, De Bock, Duysburgh, Cornelissen, et al. Thematisch rapport: SARS-CoV-2 infecties bij kinderen en jongeren van 0 tot en met 17 jaar in België, Schooljaar 2020-2021. Sciensano; 2020.

14. COVID-19 in children and the role of school settings in transmission - first update. Stockholm: European Centre for Disease Prevention and Control.; 2020 p. 59.

15. Haber NA, Clarke-Deelder E, Salomon JA, Feller A, Stuart EA. COVID-19 Policy Impact Evaluation: A guide to common design issues. ArXiv200901940 Stat [Internet]. 2021 Apr 16 [cited 2021 Sep 23]; Available from: http://arxiv.org/abs/2009.01940

16. Landeros A, Ji X, Lange K, Stutz TC, Xu J, Sehl ME, et al. An examination of school reopening strategies during the SARS-CoV-2 pandemic. PLOS ONE. 2021 May 20;16(5):e0251242.

17. Jordan I, de Sevilla MF, Fumado V, Bassat Q, Bonet-Carne E, Fortuny C, et al. Transmission of SARS-CoV-2 infection among children in summer schools applying stringent control measures in Barcelona, Spain. Clin Infect Dis [Internet]. 2021 Mar 12 [cited 2021 Aug 6];(ciab227). Available from: https://doi.org/10.1093/cid/ciab227

18. Kriemler S, Ulyte A, Ammann P, Peralta GP, Berger C, Puhan MA, et al. Surveillance of Acute SARS-CoV-2 Infections in School Children and Point-Prevalence During a Time of High Community Transmission in Switzerland. Front Pediatr [Internet]. 2021 [cited 2021 Aug 6];0. Available from: https://www.frontiersin.org/articles/10.3389/fped.2021.645577/full

19. Duysburgh E, Vermeulen M, Vandermeulen C, Roelants M, Merckx J, Desombere. Prevalence and incidence of antibodies against SARS-CoV-2 in children and school staff measured for one year in Belgium: a sero-epidemiological prospective cohort study [Internet]. 2020 [cited 2021 Oct 1]. Available from: https://www.sciensano.be/en/biblio/prevalence-and-incidence-antibodies-against-sars-cov-2-children-and-school-staff-measured-one-year

20. Statbel, het Belgische statistiekbureau | Statbel [Internet]. [cited 2021 Oct 1]. Available from: https://statbel.fgov.be/nl

21. Le portail de l’Enseignement en Fédération Wallonie-Bruxelles [Internet]. Enseignement.be. [cited 2021 Sep 29]. Available from: http://www.enseignement.be/index.php

22. Vlaams Ministerie van Onderwijs en Vorming [Internet]. [cited 2021 Sep 29]. Available from: https://www.onderwijs.vlaanderen.be/nl

23. Vlachos J, Hertegård E, B. Svaleryd H. The effects of school closures on SARS-CoV-2 among parents and teachers. Proc Natl Acad Sci. 2021 Mar 2;118(9):e2020834118.

24. Advice for the public on COVID-19 – World Health Organization [Internet]. [cited 2021 Sep 3]. Available from: https://www.who.int/emergencies/diseases/novel-coronavirus-2019/advice-for-public

25. Gettings J, Czarnik M, Morris E, Haller E, Thompson-Paul AM, Rasberry C, et al. Mask Use and Ventilation Improvements to Reduce COVID-19 Incidence in Elementary Schools - Georgia, November 16-December 11, 2020. MMWR Morb Mortal Wkly Rep. 2021 May 28;70(21):779–84.

26. Lam-Hine T. Outbreak Associated with SARS-CoV-2 B.1.617.2 (Delta) Variant in an Elementary School — Marin County, California, May–June 2021. MMWR Morb Mortal Wkly Rep [Internet]. 2021 [cited 2021 Aug 31];70. Available from: https://www.cdc.gov/mmwr/volumes/70/wr/mm7035e2.htm

27. Escandón K, Rasmussen AL, Bogoch II, Murray EJ, Escandón K, Popescu SV, et al. COVID-19 false dichotomies and a comprehensive review of the evidence regarding public health, COVID-19 symptomatology, SARS-CoV-2 transmission, mask wearing, and reinfection. BMC Infect Dis. 2021 Jul 27;21(1):710.

28. Lessler J, Grabowski MK, Grantz KH, Badillo-Goicoechea E, Metcalf CJE, Lupton-Smith C, et al. Household COVID-19 risk and in-person schooling. Science. 2021 Jun 4;372(6546):1092–7.

