## Supplements for "Measures for infection prevention and control of SARS-CoV-2 in Belgian schools between December 2020 and June 2021: a prospective cohort study"

### SUPPLEMENTARY INFORMATION

#### SUPPLEMENT I: Questionnaire M0

**COVID-19 and your school**

1. Is your school participating in this study as a primary or secondary school?
   - o Primary school
   - o Secondary school

2. Name of your school: ___________________________

3. Since reopening in May 2020, has your school been closed for any reason other than a school holiday?
   (Not including the extension of the autumn holidays 2020)
   - o YES
   - o NO

   3.1. If YES: how many days was your school closed? ________________________
   (Enter "9999" if not known)

4. Since the reopening in May 2020, have any classes been closed without the entire school being closed?
   (This means that no pupils from this particular class or classes have attended school. This does not include structural online classes)
   - o YES
   - o NO

   4.1. If YES: How many classes were closed? _________________________
   (Enter "9999" if not known)

   4.2. If YES: What was the longest closing time of a class? ______________________
   (Please enter the number of days, enter "9999" if not known)

5. Since reopening in May 2020, have any classes participating in this study been closed without the entire school being closed?
   (Classes participating in this study are: 2nd/3rd grade of primary school and 2nd grade of secondary school)
   - o YES
   - o NO

   5.1. If YES: How many of these classes were closed? _________________________
   (Enter "9999" if not known)

   5.2. If YES: What was the longest closing time of such a class?__________________________
   (Please enter the number of days, enter "9999" if not known)

6. Since reopening in May 2020, have there been any confirmed SARS-CoV-2 infection(s)* among pupils in your school?
   (*A confirmed SARS-CoV-2 infection = someone who has tested positive for a COVID-19 nasal swab test.)
   - o YES
   - o NO

   6.1. If YES, how many pupils? ____________________
   (Enter "9999" if not known)

7. Since reopening in May 2020, have there been any confirmed SARS-CoV-2 infection(s)* among teachers in your school?
   (*A confirmed SARS-CoV-2 infection = someone who has tested positive for a COVID-19 nasal swab test.)
   - o YES
   - o NO

   7.1. If YES, how many teachers? ____________________
   (Enter "9999" if not known)

8. Since reopening in May 2020, have there been any confirmed SARS-CoV-2 infection(s)* among other staff members in your school?
   (*A confirmed SARS-CoV-2 infection = someone who has tested positive for a COVID-19 nasal swab test.)
   - o YES
   - o NO

   8.1. If YES, how many staff members? ____________________
   (Enter "9999" if not known)

9. On average, how many pupils are there in a classroom?_____________________

**Infection prevention and control measures against COVID-19 at school**

10. With regard to the measures listed below, please indicate on a scale of 1 to 5 whether these measures were applied in your school or not over the period of <u>last month.</u> A score of 1 is considered as 'not applied at all' - a score of 5 is considered as 'fully applied'.

| | | 1 | 2 | 3 | 4 | 5 |
|---|---|---|---|---|---|---|
| 10.1 | The classrooms have $CO_2$ detectors (or an equivalent). | | | | | |
| 10.2 | The school has and uses a ventilation system | | | | | |
| 10.3 | Teachers are encouraged to ventilate classrooms regularly (After each class and during breaks) | | | | | |
| 10.4 | Classes take place outside as much as possible | | | | | |
| 10.5 | Breaks are spread to decrease contact between the different age groups. | | | | | |
| 10.6 | Classrooms are cleaned regularly and more frequently than previous school years (with soap and water). | | | | | |
| 10.7 | Staff rooms are cleaned regularly and more frequently than previous school years (with soap and water). | | | | | |
| 10.8 | Toilets are cleaned regularly and more frequent than previous school years (with soap and water) | | | | | |
| 10.9 | Surfaces that are touched regularly are disinfected daily | | | | | |
| 10.10 | The number of staff is limited per room. (e.g. teacher's room, staff room) | | | | | |
| 10.11 | Alcohol gel (or additional possibilities to clean hands) is made available for pupils and staff | | | | | |
| 10.12 | Pupils are given one fixed place in a fixed classroom | | | | | |
| 10.13 | Teachers change between class rooms, not the pupils | | | | | |
| 10.14 | Specific attention is paid to the strict application of safety measures in the staff room: 1) wear a mouth mask, 2) keep a distance from each other when eating or drinking, and 3) ventilate the room as much as possible. | | | | | |
| 10.15 | Lunches are taken in the classroom. If this is not possible pupils have a fixed place in the dining area | | | | | |
| 10.16 | For primary school only: distance is kept during contacts between adults. | | | | | |
| 10.17 | For primary school only: distance is kept during contacts between staff and pupils | | | | | |
| 10.18 | For primary school only: Staff wear mouth masks if the distance cannot be guaranteed. | | | | | |
| 10.19 | For secondary school only: Staff and pupils wear a mouth mask inside, even if a sufficient distance is kept. | | | | | |
| 10.20 | Secondary school only: Staff and pupils wear a mouth mask outside unless they can keep a sufficient distance. | | | | | |

**Remarks**

11. Do you have any additional remarks/comments?
   ___________________________________

#### SUPPLEMENT II: Tables

**Supplementary Table S1: Number and percentage of Dutch speaking and French speaking schools that implemented IPC measures[1] during three data collection periods (Dec 2020/Jan 2021, March 2021 and May/June 2021)[2]**

| Specific IPC measure | Dutch language network | | | French language network | | |
|---|---|---|---|---|---|---|
| | M0<br>N=44<br>n (%) | M3<br>N=44<br>n (%) | M6<br>N=45<br>n (%) | M0<br>N=39<br>n (%) | M3<br>N=38<br>n (%) | M6<br>N=37<br>n (%) |
| **IPC measures applied in both Dutch and French language networks schools (16 measures)** | | | | | | |
| *Closures (2 measures)* | | | | | | |
| School closure outside holiday breaks | 0 (0) | 4 (9) | 6 (13) | 1 (3) | 2 (5) | 2 (5) |
| Classes suspended | 20 (45) | 16 (36) | 16 (36) | 25 (64) | 16 (42) | 12 (32) |
| *Ventilation measures (4 measures)* | | | | | | |
| Classrooms have a $CO_2$ detector | 10 (23) | / | 14 (31) | 2 (5) | / | 1 (3) |
| School has and uses a ventilation system | 8 (18) | / | 5 (11) | 6 (15) | / | 4 (11) |
| Teachers are encouraged to ventilate classrooms regularly | 43 (98) | 41 (93) | 43 (96) | 37 (95) | 38 (100) | 33 (89) |
| Classes take place outside as much as possible | 2 (5) | / | 4 (9) | 3 (8) | / | 7 (19) |
| *Hygiene measures (environmental & personal) (5 measures)* | | | | | | |
| Classrooms are cleaned regularly and more frequently than previous school years | 25 (57) | 31 (70) | 36 (80) | 27 (69) | 31 (82) | 25 (68) |
| Staff rooms are cleaned regularly and more frequently than previous school years | 27 (61) | 34 (77) | 37 (82) | 26 (67) | 31 (82) | 25 (68) |
| Toilets are cleaned regularly and more frequently than previous school years | 31 (70) | 41 (93) | 39 (87) | 31 (79) | 36 (95) | 30 (81) |
| Surfaces that are touched regularly are disinfected daily | 35 (80) | 33 (75) | 34 (76) | 31 (79) | 29 (76) | 26 (70) |
| Alcohol gel (or additional possibilities to clean hands) is made available for pupils and staff | 42 (95) | 41 (93) | 43 (96) | 35 (88) | 37 (97) | 33 (89) |
| *Physical distancing measures (5 measures)* | | | | | | |
| Breaks are spread to decrease contact between different age groups | 16 (36) | 18 (41) | 15 (33) | 14 (36) | 15 (39) | 7 (19) |
| Number of staff is limited per room | 38 (86) | 36 (82) | 38 (84) | 27 (69) | 29 (76) | 26 (70) |
| Pupils have one fixed place in a fixed classroom | 31 (70) | 38 (86) | 40 (89) | 22 (56) | 23 (61) | 22 (59) |
| Teachers change between classrooms, not the pupils | 31 (70) | 35 (80) | 37 (82) | 19 (49) | 24 (63) | 18 (49) |
| Lunches are taken in the classroom. If this is not possible pupils have a fixed place in the dining area | 38 (86) | 38 (86) | 40 (89) | 32 (82) | 38 (100) | 28 (76) |

[1] All measures, except for closures, are scored on a scale ranging from 1 (not applied at all) to 5 (fully applied). Measures with a score of '4' or '5' are considered 'implemented' by the school.

[2]Schools who did answer to certain measures were considered to not apply these measures. The number of missing schools is shown in Table S3.

IPC: infection prevention and control; M0: data collection period at study month 0, 3 Dec 2020 - 28 Jan 2021; M3: data collection period at study month 3, 1 - 26 Mar 2021; M6: data collection period at study month 6, 17 May – 11 Jun 2021; n (%): absolute number of schools (percentage of schools) that implemented the measure; N: number of schools that completed the questionnaire.

**Supplementary Table S2: Number and percentage of missing values regarding IPC measure implementation in Belgian primary and secondary schools during three data collection periods (Dec 2020/Jan 2021, March 2021 and May/Jun 2021)**

| Specific IPC measure | Primary schools | | | Secondary schools | | |
|---|---|---|---|---|---|---|
| | M0<br>N=43<br>n (%) | M3<br>N = 44<br>n (%) | M6<br>N = 43<br>n (%) | M0<br>N=40<br>n (%) | M3<br>N=38<br>n (%) | M6<br>N=39<br>n (%) |
| **IPC measures applied in both primary and secondary schools** | | | | | | |
| Ventilation measures (4 measures) | | | | | | |
| Classrooms have a $CO_2$ detector | 3 (7) | / | 6 (14) | 1 (3) | / | 6 (15) |
| School has and uses a ventilation system | 2 (5) | / | 9 (21) | 6 (15) | / | 3 (8) |
| Teachers are encouraged to ventilate classrooms regularly | 0 (0) | 0 (0) | 1 (2) | 1 (3) | 0 (0) | 3 (8) |
| Classes take place outside as much as possible | 0 (0) | / | 1 (2) | 1 (3) | / | 4 (10) |
| Hygiene measures (environmental & personal) (5 measures) | | | | | | |
| Classrooms are cleaned regularly and more frequently than previous school years | 1 (2) | 0 (0) | 2 (5) | 3 (8) | 0 (0) | 4 (10) |
| Staff rooms are cleaned regularly and more frequently than previous school years | 0 (0) | 0 (0) | 2 (5) | 2 (5) | 0 (0) | 3 (8) |
| Toilets are cleaned regularly and more frequently than previous school years | 0 (0) | 1 (2) | 1 (2) | 2 (5) | 0 (0) | 3 (8) |
| Surfaces that are touched regularly are disinfected daily | 1 (2) | 0 (0) | 1 (2) | 2 (5) | 1 (3) | 3 (8) |
| Alcohol gel (or additional possibilities to clean hands) is made available for pupils and staff | 1 (2) | 0 (0) | 2 (5) | 2 (5) | 0 (0) | 3 (8) |
| Physical distancing measures (5 measures) | | | | | | |
| Breaks are spread to decrease contact between different age groups | 1 (2) | 2 (5) | 4 (9) | 1 (3) | 1 (3) | 4 (10) |
| Number of staff is limited per room | 3 (7) | 0 (0) | 2 (5) | 2 (5) | 1 (3) | 3 (8) |
| Pupils have one fixed place in a fixed classroom | 1 (2) | 0 (0) | 2 (5) | 3 (8) | 0 (0) | 3 (8) |
| Teachers change between classrooms, not the pupils | 6 (14) | 5 (11) | 8 (19) | 2 (5) | 0 (0) | 4 (10) |
| Lunches are taken in the classroom. If this is not possible pupils have a fixed place in the dining area | 3 (7) | 0 (0) | 1 (2) | 3 (8) | 0 (0) | 4 (10) |
| **IPC measures applied in primary schools only (3 measures)** | | | | | | |
| Staff wear a mask if sufficient distance cannot be maintained | 2 (5) | 0 (0) | 1 (2) | | | |
| Distance is kept during contacts between adults | 1 (2) | 0 (0) | 1 (2) | | | |
| Distance is kept during in contacts between staff and pupils | 4 (9) | 0 (0) | 1 (2) | | | |
| **IPC measures applied in secondary schools only (2 measures)** | | | | | | |
| Secondary schools only: Staff and pupils always wear a mask inside | | | | 3 (8) | 1 (3) | 6 (15) |
| Secondary schools only: Staff and pupils wear a mask outside unless they can keep sufficient distance | | | | 3 (8) | 1 (3) | 5 (13) |

IPC: infection prevention and control; M0: data collection period at study month 0, 3 Dec 2020 - 28 Jan 2021; M3: data collection period at study month 3, 1 - 26 Mar 2021; M6: data collection period at study month 6, 17 May – 11 Jun 2021; n (%): absolute number of schools (percentage of schools) that implemented the measure; N: number of schools that completed the questionnaire.

**Supplementary Table S3 : Number and percentage of the missing values regarding IPC measure implementation in schools of the Dutch and French language network during three data collection periods (Dec 2020/Jan 2021, March 2021 and May/Jun 2021)**

| Specific IPC measure | Dutch language network | | | French language network | | |
|---|---|---|---|---|---|---|
| | M0<br>N=44<br>n (%) | M3<br>N=44<br>n (%) | M6<br>N=45<br>n (%) | M0<br>N=39<br>n (%) | M3<br>N=38<br>n (%) | M6<br>N=37<br>n (%) |
| **IPC measures applied in both Dutch and French language networks schools (16 measures)** | | | | | | |
| *Ventilation measures (4 measures)* | | | | | | |
| Classrooms have a $CO_2$ detector | 1 (2) | / | 4 (9) | 3 (8) | / | 8 (22) |
| School has and uses a ventilation system | 1 (2) | / | 3 (7) | 7 (18) | / | 9 (24) |
| Teachers are encouraged to ventilate classrooms regularly | 1 (2) | 0 (0) | 1 (2) | 0 (0) | 0 (0) | 3 (8) |
| Classes take place outside as much as possible | 0 (0) | / | 1 (2) | 1 (3) | / | 4 (11) |
| Hygiene measures (environmental & personal) (5 measures) | | | | | | |
| Classrooms are cleaned regularly and more frequently than previous school years | 4 (9) | 0 (0) | 1 (2) | 0 (0) | 0 (0) | 4 (11) |
| Staff rooms are cleaned regularly and more frequently than previous school years | 2 (5) | 0 (0) | 1 (2) | 0 (0) | 0 (0) | 4 (11) |
| Toilets are cleaned regularly and more frequently than previous school years | 2 (5) | 0 (0) | 1 (2) | 1 (3) | 1 (3) | 3 (8) |
| Surfaces that are touched regularly are disinfected daily | 3 (7) | 1 (2) | 1 (2) | 0 (0) | 0 (0) | 3 (8) |
| Alcohol gel (or additional possibilities to clean hands) is made available for pupils and staff | 2 (5) | 0 (0) | 1 (2) | 0 (0) | 0 (0) | 4 (11) |
| Physical distancing measures (5 measures) | | | | | | |
| Breaks are spread to decrease contact between different age groups | 1 (2) | 0 (0) | 1 (2) | 1 (3) | 3 (8) | 7 (19) |
| Number of staff is limited per room | 3 (7) | 0 (0) | 1 (2) | 2 (5) | 1 (3) | 4 (11) |
| Pupils have one fixed place in a fixed classroom | 2 (5) | 0 (0) | 2 (4) | 2 (5) | 0 (0) | 3 (8) |
| Teachers change between classrooms, not the pupils | 3 (7) | 3 (7) | 4 (9) | 5 (13) | 2 (5) | 8 (22) |
| Lunches are taken in the classroom. If this is not possible pupils have a fixed place in the dining area | 4 (9) | 0 (0) | 1 (2) | 2 (5) | 0 (0) | 4 (11) |

IPC: infection prevention and control; M0: data collection period at study month 0, 3 Dec 2020 - 28 Jan 2021; M3: data collection period at study month 3, 1 - 26 Mar 2021; M6: data collection period at study month 6, 17 May – 11 Jun 2021; n (%): absolute number of schools (percentage of schools) that implemented the measure; N: number of schools that completed the questionnaire.
